# Predicting Physiological Response in Heart Failure Management: A Graph Representation Learning Approach using Electronic Health Records

**DOI:** 10.1101/2023.01.27.23285129

**Authors:** Shaika Chowdhury, Yongbin Chen, Andrew Wen, Xiao Ma, Qiying Dai, Yue Yu, Sunyang Fu, Xiaoqian Jiang, Nansu Zong

## Abstract

Heart failure management is challenging due to the complex and heterogenous nature of its pathophysiology which makes the conventional treatments based on the “one size fits all” ideology not suitable. Coupling the longitudinal medical data with novel deep learning and network-based analytics will enable identifying the distinct patient phenotypic characteristics to help individualize the treatment regimen through the accurate prediction of the physiological response. In this study, we develop a graph representation learning framework that integrates the heterogeneous clinical events in the electronic health records (EHR) as graph format data, in which the patient-specific patterns and features are naturally infused for personalized predictions of lab test response. The framework includes a novel Graph Transformer Network that is equipped with a self-attention mechanism to model the underlying spatial interdependencies among the clinical events characterizing the cardiac physiological interactions in the heart failure treatment and a graph neural network (GNN) layer to incorporate the explicit temporality of each clinical event, that would help summarize the therapeutic effects induced on the physiological variables, and subsequently on the patient’s health status as the heart failure condition progresses over time. We introduce a global attention mask that is computed based on event co-occurrences and is aggregated across all patient records to enhance the guidance of neighbor selection in graph representation learning. We test the feasibility of our model through detailed quantitative and qualitative evaluations on observational EHR data.

## Introduction

Heart failure (HF) is a complex clinical syndrome resulting from either structural or functional cardiac impairment in the capacity of ventricles to fill up with or eject blood^1^ and is associated with significant morbidity, mortality and health care expenditures worldwide^2,3^. Heart failure is not a singular disease but is rather characterized by a broad spectrum of etiologies and pathophysiologies leading to heterogeneous patient subgroups^3,4^. This phenotypic diversity ensues variability in the treatment outcomes across patients, thus imposing a great challenge on effective intervention administration in curing heart failure.

The key to resolving this disease heterogeneity is in identifying the patient subgroups underlying the physiological deviations (i.e., phenotypes)^5,6,7^. This notion intuitively portrays the real-world clinical prognosis workflow – the physician first performs diagnostic tests to quantify the phenotypical observations related to the patient that would help them make a potential diagnosis^8^ and then tracks the disease prognosis through the patient’s response to treatment. The conventional approaches to heart failure management, however, have been inadequate in contemplating the phenotypic heterogeneity of this complex disease as treatment is extrapolated based on the average population, inducing suboptimal patient care and quality of life. Apparently, heart failure has the prospect of benefitting from stratified management strategies (i.e., precision medicine) that would ensure targeted treatment and prevention for each heart failure subgroup, while considering the individual differences among patients.

Although the general focus of precision medicine has been on omics-type “big data”, in particular genomics data, nevertheless, in the case of heart failure the genomic-centric approach is not ideal owing to its limited genetic components and associated environmental triggers in most instances^7,9^. In the recent past, Electronic Health Records (EHR) have contributed to generating enormous volumes of time-based phenotypic data that is characterized as intrinsically “big” due to its complexity (i.e., variety) and the bulk of heterogeneous information available per patient (i.e., volume), that is much greater in amount compared to any other patient databases^10^. The power of precision medicine lies in sieving through this EHR data to mine the patients’ health-related patterns and features that would enable the stratification of the heart failure cohort into therapeutically homogeneous patient subgroups. In order to make sense of this longitudinal data and successfully establish the patient patterns into actionable insights, it is of critical importance to harness advanced analytics such as deep learning for improved prognostication of treatment outcomes.

In physiological response prediction, regression analysis on a biomarker is performed as an indicator of the patient’s pharmacological response to a therapeutic intervention. If there exists a close association between a biomarker and a hard clinical endpoint (e.g., mortality, hospitalization) reflected through the changes in the biomarker measurements following treatment, it suffices to substitute the hard end point with the biomarker as a surrogate endpoint^11^. Blood pressure (BP) provides a non-invasive measurement of cardiac function and as supported by several studies, serves as a physiological biomarker that has been shown to have a consistent relationship with cardiovascular mortality and morbidity^12,13^. According to large cohort studies and randomized controlled trials, blood pressure is regarded as a valid surrogate endpoint as high blood pressure was found to be a risk factor for cardiovascular events, with a reduced level of blood pressure diluting the risk of such adverse outcomes^12,14,15,16^. Therefore, predicting the prognostic value of blood pressure as the drug response could possibly uncover the differences in the pathophysiological mechanisms defining the heterogeneous prognosis of heart failure to help guide the appropriate therapies to the patient subgroups; thus could serve as a valuable tool to cross-check the physician’s decision making in the intervention administration. The adoption of computer-assisted outcome prediction in the form of deep learning models holds great promise in providing sufficient computational and statistical power to understand and interpret the role of biomarkers in deriving prognostic insights and identifying the phenotypes towards enhancing tailored therapeutic strategies in heart failure management.

In spite of the fact that traditional deep learning models such as multilayer perceptron (MLP), convolutional neural network (CNN) and recurrent neural network (RNN) have yielded remarkable performance in treatment outcome prediction tasks^17-21^, they fail to embody the complex topological structure of the non-euclidean data^22,23^. The physiological lab measurements in EHR form multivariate time-series data, which is present in a non-euclidean space as defined by the temporal and spatial dependencies^24,25^ among the clinical events. On one hand, the sequential measurements recorded over different visits for each clinical event could evolve over time to accurately monitor heart failure severity and progression, manifesting an inherent temporality in EHR. On the other hand, the synergistic interactions among different clinical events in the causal pathway of heart failure pathophysiology portray the spatial dynamics. This spatial-temporal structure of physiological recordings in EHR exists as an irregular grid due to the diverse and arbitrary linkages among the clinical events, which can be naturally formalized as graph data. Generalizing deep learning on graph-structured data offers the combined benefit of harnessing the data-driven capability of deep learning techniques to effectively model the intrinsic relationships among the nodes in the graph. Graph representation learning is such a paradigm that encodes the graph through projection to a low-dimensional vector space while maximally preserving the graph topology and node properties and has witnessed enormous success in various biomedical applications^23,26^. The utility of graph representation learning in treatment outcome prediction is currently in its infancy. A recent work^27^ performed lab test response prediction by first using Transformers to encode the longitudinal diagnosis and medication information in the patient’s EHR. It then uses Graph Attention Networks (GAT) to encode the similarity among the patients and the lab interaction-based external knowledge. The representations are finally concatenated together with the patient’s past lab test response information to get the patient representation. However, a major limitation of this work is that the Transformer-encoded sequential representation and the GAT-encoded graph representation are learned separately and then combined, which could cause important information loss along the spatial domain.

To directly forecast the changing of the physiological biomarker which is critical to facilitate physicians in decision-making for HF patients, we propose an end-to-end graph-based unified framework that learns the patient representation by jointly modeling the underlying spatial and temporal patterns in the EHR and optimizes it for blood pressure forecast. First, to model the historical physiological information in the patient’s EHR, we construct a knowledge graph relating the heterogeneous clinical events in the medical history through temporal connectivity and timestamp features. We then propose a Transformer-based Graph Neural Network model to propagate and exchange patient-specific information across neighboring nodes to simultaneously learn the spatial and temporal interactions in the graph structure to support personalized response predictions.

## Methods

### Data Collection and Preparation

The study cohort was obtained from Mayo Clinic’s United Data Platform (UDP), a data warehouse that contains, consolidates, and standardizes all clinical data collected within the institution. We identify patients with Heart Failure conditions using the diagnosis codes listed in Table 1. With each patient record corresponding to a single visit, we utilize the demographics, diagnosis, lab test and medication information to create the HF dataset. We evaluate the pharmacological effect of five categories/classes of drugs - *Angiotensin-converting-enzyme inhibitors* (ACEI), *Beta Blocker* (BB), *Angiotensin II receptor blockers (*ARB), *Statin* and *Loop Diuretic* (LD) - and create the dataset for each separately by using the corresponding medication codes to retrieve the relevant patient records. Refer to Table 2 for the medications belonging to each category and Table 3 for the data statistics per category. We use *N*_*pat*_ in the rest of the paper to denote the total number of patients in each dataset.

**Table 1.**
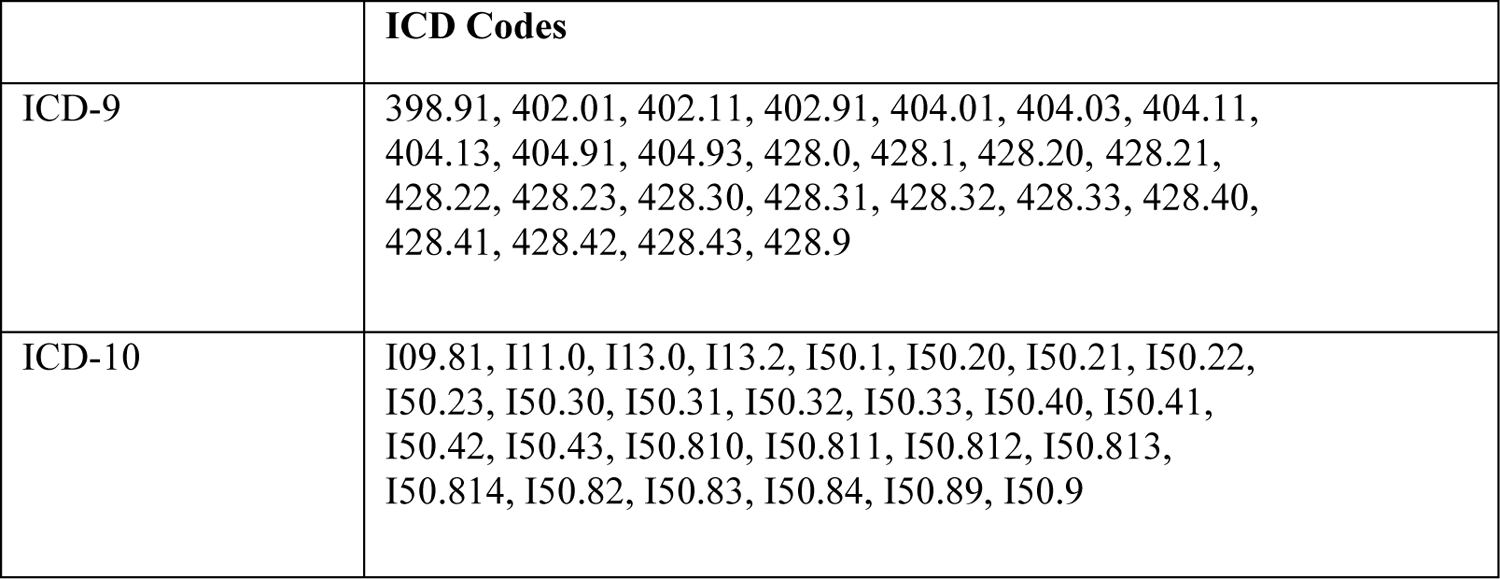
Diagnosis codes associated with HF

|  | ICD Codes |
| --- | --- |
| ICD-9 | 398.91, 402.01, 402.11, 402.91, 404.01, 404.03, 404.11, 404.13, 404.91, 404.93, 428.0, 428.1, 428.20, 428.21, 428.22, 428.23, 428.30, 428.31, 428.32, 428.33, 428.40, 428.41, 428.42, 428.43, 428.9 |
| ICD-10 | I09.81, I11.0, I13.0, I13.2, I50.1, I50.20, I50.21, I50.22, I50.23, I50.30, I50.31, I50.32, I50.33, I50.40, I50.41, I50.42, I50.43, I50.810, I50.811, I50.812, I50.813, I50.814, I50.82, I50.83, I50.84, I50.89, I50.9 |

**Table 2.** HF drug category and medications

| Category | Medication Name |
| --- | --- |
| ACEI | Benazepril, Lotensin, Captopril, Enalapril, Vasotec, Fosinopril, Lisinopril, Prinivil, Zestril, Moexipril, Perindopril, Quinapril, Accupril, Ramipril, Altace, Trandolapril |
| Beta Blocker | Acebutolol, Atenolol, Tenormin, Bisoprolol, Zebeta, Metoprolol, Lopressor, Toprol XL, Nadolol, Corgard, Nebivolol, Bystolic, Propranolol, Inderal, InnoPran XL |
| ARB | Azilsartan, Edarbi, Candesartan, Atacand, Eprosartan, Irbesartan, Avapro, Losartan, Cozaar, Olmesartan, Benicar, Telmisartan, Micardis, Valsartan, Diovan |
| Statin | Atorvastatin, Lipitor, Lovastatin, Altoprev, Pitavastatin, Livalo, Zypitamag, Pravastatin, Pravachol, Rosuvastatin, Crestor, Ezallor, Simvastatin, Zocor |
| Loop Diuretic | Chlorothiazide, Chlorthalidone, Hydrochlorothiazide, Indapamide, Metolazone, Bumetanide, Bumex, Ethacrynic acid, Edecrin, Furosemide, Lasix, Torsemide, Soaanz, Amiloride, Midamor, Eplerenone, Inspra, Spironolactone, Aldactone, Carospir, Triamterene, Dyrenium |

**Table 3.**
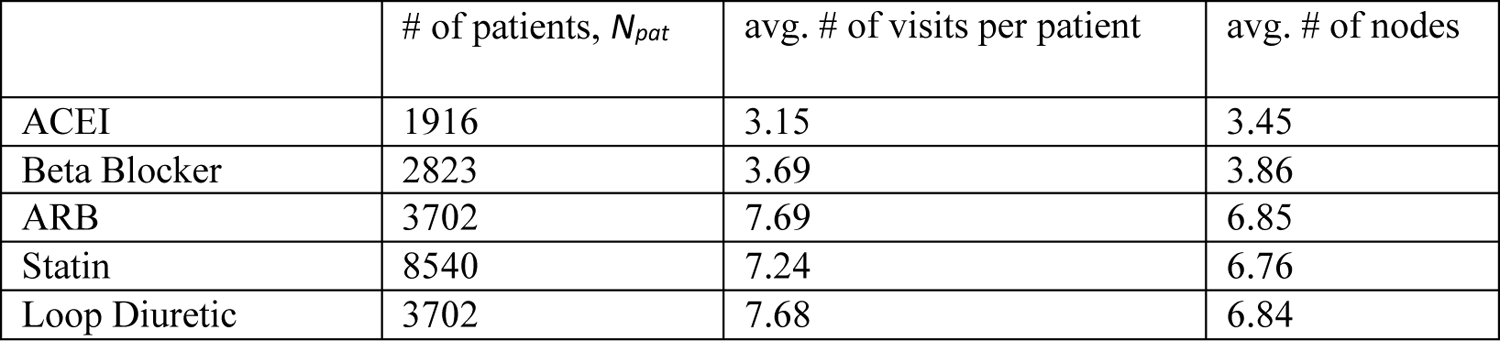
Statistics of the datasets for the five HF drug categories

| | # of patients, $N_{pat}$ | avg. # of visits per patient | avg. # of nodes |
| --- | --- | --- | --- |
| ACEI | 1916 | 3.15 | 3.45 |
| Beta Blocker | 2823 | 3.69 | 3.86 |
| ARB | 3702 | 7.69 | 6.85 |
| Statin | 8540 | 7.24 | 6.76 |
| Loop Diuretic | 3702 | 7.68 | 6.84 |

### Problem Statement

In this retrospective observational study, we predict the patient’s drug response as measured by the BP lab test (i.e., DBP) based on the longitudinal patient history in EHR. The patient history can be perceived as a collection of EHR records associated with heart failure conditions that can be represented as a sequence of time-ordered visits, while each visit is comprised of a list of clinical concepts essentially summarizing the prognostic and interventional events involved in heart failure management. Formally, let P = (V_1_, V_2_, …., V_T_) denote the EHR records of a single patient with total T visits, where V_i_ = (c_1_^i^, c_2_^i^, …., c_|_v_i|_^i^) is a visit in P arranged by the time of occurrence and c_j_^i^ = (e_j_^i^, v_j_^i^, t_j_^i^) is a clinical event in V_i_ composed of a tuple of the type of event e_j_^i^ ϵ E, the observed value of the event v_j_ ^i^ and the timestamp of the event t_j_^i^ ϵ R^*^_+_. Here, E corresponds to the unique set of clinical events, v_j_ ^i^ is either a categorical value or a numerical measurement depending on the type of event and R^*^_+_ is the set of positive real numbers. Given the patient’s EHR sequence P containing the time-varying heterogeneous phenotypic events from E, the goal of this study is to forecast the value of the hemodynamic event BP lab test, Ŷ, in the future time step (i.e., visit) via learning a graph-based mapping function *f*: P → Ŷ.

### Graph Construction

In order to mimic the intricacies of the patient’s complex treatment process, we create a distinct health network for each patient by transforming the patient-specific events in the EHR sequence P to a knowledge graph G = (V, E, A, X), where V is the set of vertices, E is the set of edges connecting the vertices, A is the adjacency matrix and X is the node feature matrix. We first consider clinical events derived from three data sources in EHR to form the nodes V in G: demographics *Demographics* ϵ {age, gender}, lab tests *Lab* ϵ {DBP, SBP, SVR} and diagnosis *Comorbidity* ϵ {hypertension, hyperlipidemia, shortness of breath, atrial fibrillation, cancer, diabetes mellitus, dyslipidemia}. Here, DBP, SBP and SVR refer to the hemodynamic variables, diastolic blood pressure, systolic blood pressure, and systematic vascular resistance respectively, and Comorbidity denotes the seven comorbidities with the highest frequency in our dataset. We propose graph construction from two different perspectives – *single data source* and *multi-data source* – so as to assess the individual and collaborative informativeness of the data sources in predicting HF treatment outcomes. That is, each data source is leveraged in isolation for single-data source graph construction, while all the variables across the three data sources contribute as nodes in the multi-data source graph, as depicted in Figure 1. To account for the spatial and temporal dependencies within the multivariate physiological recordings in EHR, we define nodes with respect to each event as well as the chronology of the event. The chronology of the event signifies how the value of the phenotypic variable varies over the visits. Formally speaking, given the sequential observations associated with a particular clinical event C = {c_t_ | t ϵ {1,2,..T}, c_t_ ϵ Demographics or c_t_ ϵ Lab or c_t_ ϵ Comorbidity} for the single data source and C = {c_t_ | t ϵ {1,2,..T}, c_t_ ϵ Demographics U c_t_ ϵ Lab U c_t_ ϵ Comorbidity} for multi-data source, where T denotes the total visits in the patient’s EHR and U is the union operation, we introduce a new node in G for the event at each time step t, as depicted in Figure 1. We further explicitly incorporate the temporal aspect of EHR such that two clinical events form an edge e ϵ E if they appear consecutively in the time-ordered sequence associated with the event. However, instead of considering the direct future event as the only neighbor in accordance with the temporal directionality of the sequence, we include all the future events as the one-hop neighborhood to capture long-term dependencies. Additionally, we embrace an undirected topology for the sake of bidirectional propagation of the past and future event information, endorsed based on less favorable preliminary results with directed connectivity and previous findings^28,29^. The adjacency matrix A ϵ R^|V| x |V|^ summarizes this temporal graph structure knowledge whereby its (i,j)-th entry is 1 if e(i,j) ϵ E, otherwise it is a 0. From the aforementioned, without loss of generality, recall that each event can be defined as a tuple c_t_ = (e_t_, v_t_, t_t_). We use this nuanced information to annotate event-specific features in the graph. First, we designate the event type e_t_ (e.g., SBP) as the node name and subscript it by the time step it occurred in. Second, the event value v_t_ is assigned as the node feature, indicative of the patient’s prognostic state. Specifically, for the variables in the data sources Demographics and Lab we consider the raw EHR features and then apply MinMax normalization. While for Comorbidity, we represent the node feature by applying the Term Frequency-Inverse Document Frequency (Tf-idf). Since the base model of the proposed approach in this study is a Transformer which by design is invariant to sequence order^30^, we draw on the timestamp of the event t_t_ to infuse positional information into the graph structure. The timestamp is normalized and added to the event value v_t_ as the position-aware node feature matrix X ϵ R^|V| x 1^. Note that any missing time-stamped events in the patient’s EHR will not appear in the graph G. As a result, mapping the original multivariate EHR sequence to a graph structure facilitates in seamlessly tackling the prevalent missingness issue surrounding multivariate sequence problems without having to resort to data imputation.

**Figure 1.**
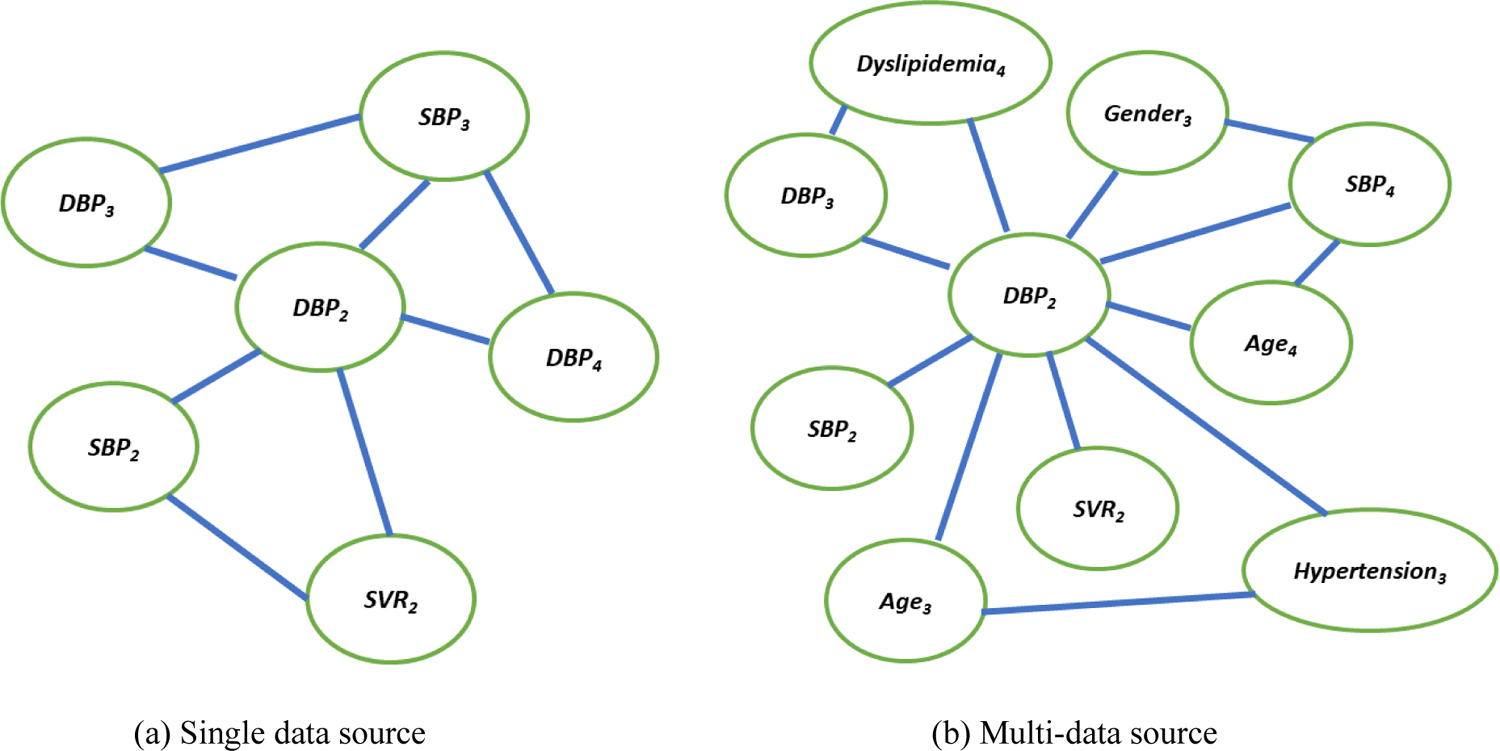
Graph Construction. Note that the exhaustive connections are not displayed.

### Model Overview

With the constructed patient knowledge graph, we then customize the Graph Neural Network (GNN) model to map the health network to a low dimensional vector, that would encapsulate the patient-specific phenotypic features discriminative for personalized decision making. The vanilla GNN relies on message passing for node representation learning by iteratively propagating and gathering messages from adjacent nodes (i.e., neighborhood aggregation step), then using this information alongside its own features to refine its representation (i.e., updating step). However, a GNN considers all the neighbors to equally contribute to the representation update, which could downplay the actual significance of some variables in relation to the clinical outcome. We compensate for this limitation by exploiting the Transformer’s self-attention mechanism to prioritize the neighborhood. Provided the input feature matrix *X* of all the nodes in G, self-attention first projects it to the query, key and value spaces, *Q* = *XW*_q_, *K* = *XW*_k_, *V* = *XW*_v_, using the trainable weight matrices *W*_q_, *W*_k_ and *W*_V_ respectively. Then scaled dot product is employed to compute the attention as,

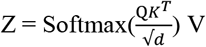

where d is the dimensionality of the attention head and is used in the scaling factor for numerical stability. Here, Z ϵ R^|V| x d^ holds the output node representations generated by the self-attention as the weighted sum of the linearly transformed input nodes’ features. In addition, Transformer’s self-attention repeats this mechanism in parallel several times (i.e., multi-head attention) to jointly learn from different representation subspaces. We call this model a *Graph Transformer*.

The original Transformer has memory and computation overhead quadratic to the graph cardinality |V|, which could be problematic in training larger patient knowledge graphs, so we adopt a sampling strategy^30^. Contrary to computing the pairwise attention score with respect to every node n ϵ V, we only sample a subset of the nodes as the neighborhood for each node and feed it as the input matrix into the self-attention component.

The self-attention is crucial for physiological response prediction as it is a key component in accurately modeling the implicit structure of the latent spatial relationships among the clinical events in EHR. However, recall that there is also an explicit structure of the input graph G described by the adjacency matrix A which stores the temporal event connections. As the Transformer naturally assumes any graph as fully connected, self-attention would not be able to recognize this explicit temporal structure. To tackle this, we augment a GNN layer on top of the Graph Transformer, which models the temporality inductive bias by inputting the adjacency matrix A. This way the proposed framework is capable of embedding the spatial-temporal patterns in G at once.

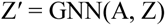

Finally, we readout the output node representations Z′ into a single vector representing the patient’s health profile by summation. This graph vector is then passed through a linear layer to forecast the BP level in the future visit, Ŷ.

### Global Attention Mask

With the aim of stratifying HF treatment into homogeneous patient subgroups with predictable responses, we synthesize the distinct patient knowledge graph with prognostic patterns analyzed across the entire HF cohort through the guidance of a *global attention mask*. The influence of global dependencies in the outcome prediction will ensure that the patient similarities across the phenotypic spectrum are assimilated with their individual clinical variations to realize more informed HF management. The global attention mask achieves this by building a binary event co-occurrence matrix, M ϵ R^|V| x |V|^, drawn from all the patients’ records in the EHR. This is to say, if two clinical events appear together in any record, we set the corresponding entry in M to 1, otherwise, it is set to 0. We then use this event co-occurrence matrix to redefine self-attention with the attention mask function *Mask*, as notated below,

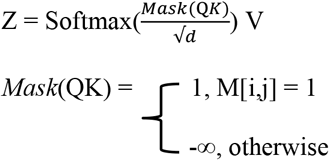

where i and j are the positions in the query and key respectively. Concretely, this mask function guides the selection of neighbors for attention computation by allowing only the co-occurred events to be attended, else ignoring the input position. This way, a more robust node representation is learned based on the knowledge aggregated from both the patient’s and other patients’ EHR profiles, possibly suggestive of actual physiological correlations in HF pathophysiology, rather than merely relying on randomly sampled nodes as neighbors.

### Optimization and Evaluation Metrics

Given the ground truth BP measurement recorded in the last visit of the patient, Ƴ, we use the mean squared error (MSE) as the objective function:

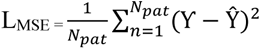

We use Adam^32^ optimizer to minimize the loss function.

We evaluate the effectiveness of the model using the mean absolute error (MAE), mean squared error (MSE) and root mean squared error (RMSE), with computations as below,

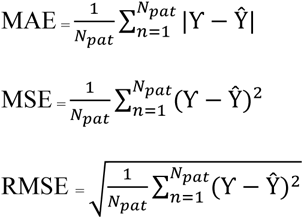

#### Experimental Setup

For the model evaluation, we adopt the 10-fold cross validation technique. In every fold, 9 equal-sized disjoint subsets are trained for 50 epochs and tested on the remaining held-out subset. The average of the performances on the 10 held-out subsets is reported as the model’s final prediction performance. We set the batch size during training to 4 and use a learning rate of 5e^-4^.

#### Experiments

We divide the conducted experiments into three parts to investigate the graph-based framework’s drug response prediction performance in HF treatment from a holistic perspective. In the first part, we focus on the proposed model’s design and demonstrate its capability through comparisons on four grounds – data source, baseline models, ablation study and the number of steps to forecast. In the second part, we assess the model’s predictive power on the individual drug categories and their combinations besides. In the last part, we shed light on the treatment response differences between the HF subtypes quantitatively and qualitatively. For the set of experiments in Part 1 Evaluation, we assess only the ACEI medications as the representative drug category as Part 2 Evaluation covers the comparisons among all the drug categories.

### Part 1 Evaluation

The data source corresponds to the type of input data from EHR used to construct the patient knowledge graph and is an important evaluation criterion as it could elucidate insights into the source-specific clinical events’ contributions in the drug response characterizing the patient profiles in HF treatment. Figures 2 - 4 top left subplots report this performance comparison between the three single data sources – Demographics, Lab and Comorbidity – which includes the subset of variables specific to the data source as the predictors, and the multi-data source (i.e., ALL) which includes all the variables. Note that for the single data sources we also incorporate the DBP variables in the past visits for the graph construction as it is predicted as the outcome of interest. Among the three single data sources, Lab performs the best across all the metrics followed by Comorbidity, with Demographics performing the worst. The Lab tests DBP, SBP and SVR are considered as risk factors imperative in HF prognosis^33^ and are routinely monitored as part of the EHR, so the good results are not surprising. The HF cohort predominantly consists of older patients (e.g., 74 was the most prevalent age in our ACEI cohort), who are also more likely to have multiple comorbidities^34^, such as hypertension, diabetes mellitus, atrial fibrillation, and hyperlipidemia, which further contribute to the heterogeneity of HF^35^. So, using the Comorbidity information is beneficial as indicated by its satisfactory performance and identifying the combinations of the comorbidities corresponding to the different phenogroups as a next step could lead to targeted HF treatment. Most Demographics information include static variables (e.g., gender) which could remain time-invariant throughout the treatment course and hence do not provide discriminative features in the temporal modeling of drug response prediction. This could attribute to the Demographics data source performing the worst. Generally, the single data sources are seen to perform better than the multi-data source (ALL), with a performance gap of around 4.9% in RMSE against the best-performing data source Lab. For the subsequent performance comparisons, we use the best-performing data source, Lab, as the input data.

**Figure 2.**
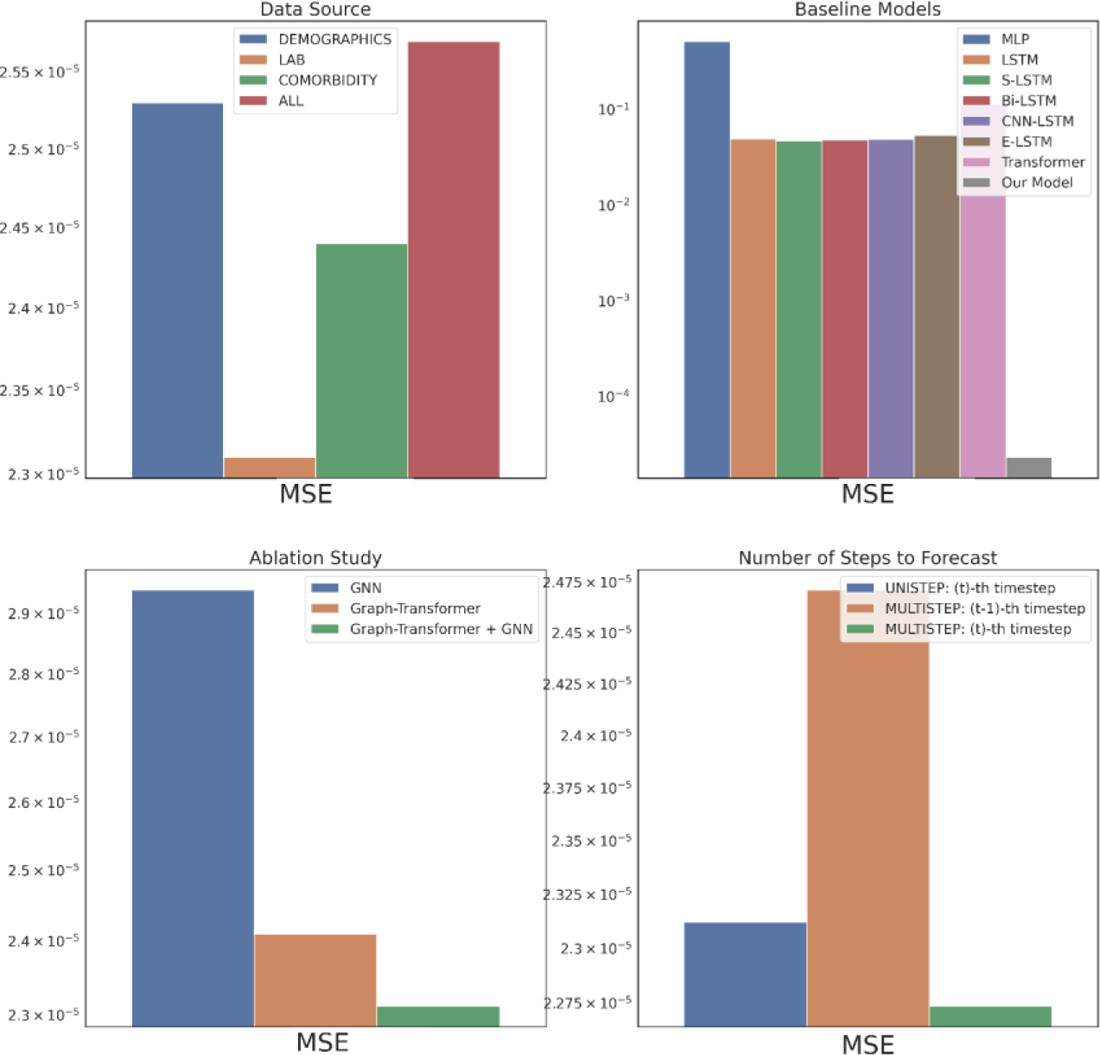
Part 1 Evaluation in MSE

**Figure 3.**
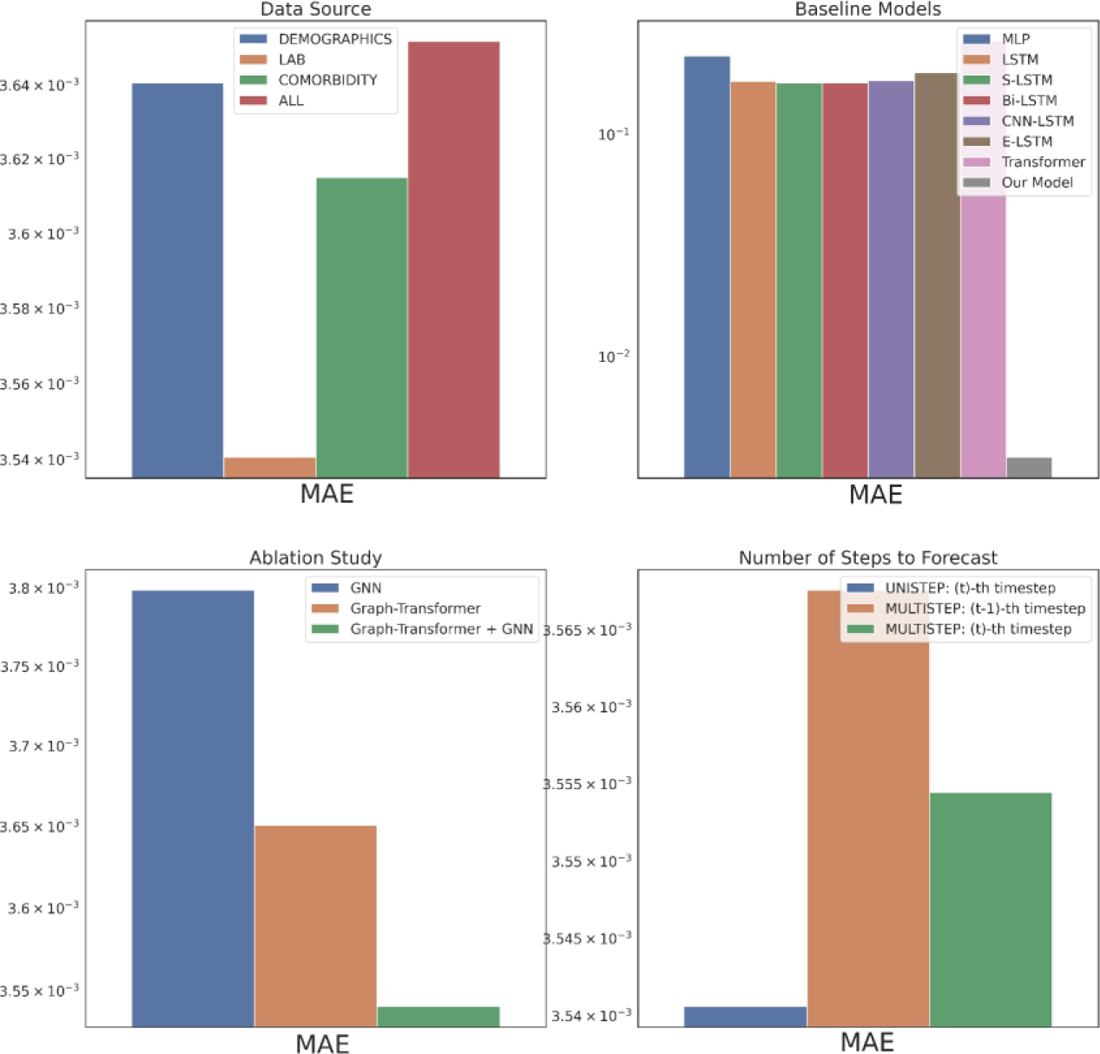
Part 1 Evaluation in MAE

**Figure 4.**
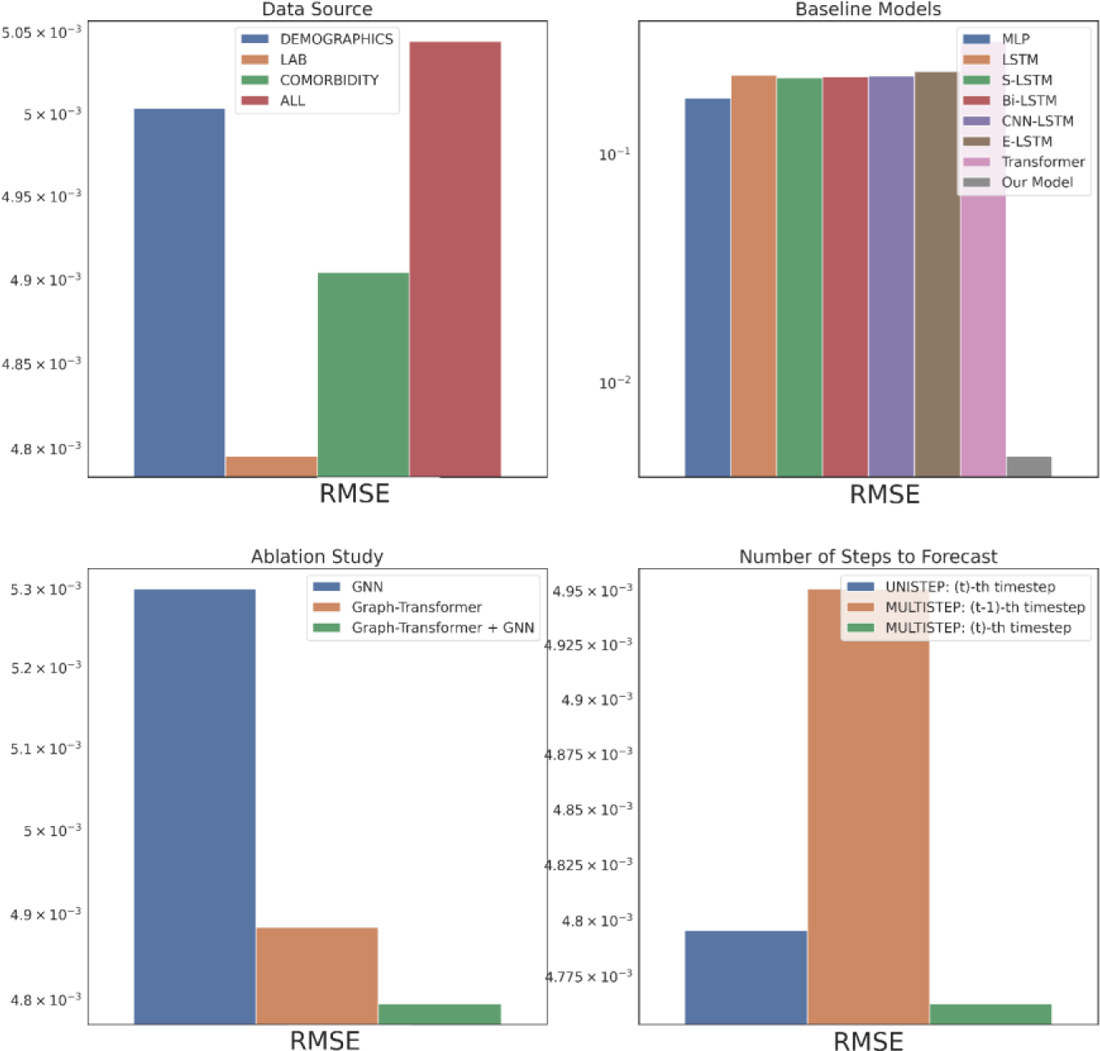
Part 1 Evaluation in RMSE

We compare the proposed model’s performance with the following deep learning models as the baselines: multi-layer perceptron (MLP), long short-term memory model (LSTM)^36^, stacked LSTM (S-LSTM) with 2 LSTM layers, bidirectional LSTM (Bi-LSTM)^37^, CNN-LSTM consisting of a CNN layer^38^ and an LSTM layer, ensemble LSTM (E-LSTM) that combines predictions based on Stacked Generalization^39^ and Transformer^40^, depicted in Figures 2-4 top right subplots. Overall, our proposed model is able to consistently outperform all the baselines significantly. This corroborates representing the multivariate physiological findings in EHR as graph-structured data, as otherwise all the baselines directly used the sequential information and failed to effectively model the per-variable (i.e., temporal) and inter-sequence (i.e., spatial) dependencies.

An ablation study is carried out to verify the impact of the proposed model’s components, GNN and Graph Transformer, as depicted in Figures 2-4 bottom left subplots. Individually, the GNN and Graph Transformer models underperform in comparison to the complete model Graph Transformer + GNN as empirically the complete model is able to achieve a 9.5% and 1.8% RMSE reductions over the GNN and Graph Transformer respectively. In the case of GNN, this performance decline possibly arises from its failure to distill the important features because of equal weighting during node update. Although the Graph Transformer addresses this limitation by soft-selecting the neighbors, it loses on the explicit graph structure that defines the temporal relationships between the clinical events. Hence, incrementally building on both components to get the complete model leads to the best performance.

In the original problem setting, our model forecasts the drug response in the patient’s last visit having trained on all the previous visits’ EHR information. Practically in the actual clinical scenario, however, a physician would benefit more from knowing the treatment effects on the patient for multiple visits to be able to intervene in advance and prevent any negative clinical outcomes. Figures 2-4 bottom right subplots illustrate the model’s performance in this phenomenon (i.e., MULTISTEP) for the last two visits (t^-th^ and (t-1)^-th^ timesteps) and contrasts it with its original performance in the last visit (i.e., UNISTEP). The results show comparable performance for the UNISTEP and MULTISTEP’s t^-th^ prediction but degrades in the MULTISTEP’S (t-1)^-th^ timestep.

### Part 2 Evaluation

In the second set of experiments, we analyze the treatment response by tapping into the medication information from five different angles – drug category, top drug per category, drug combinations within a category (intra-category), drug combinations between two categories (inter-category) and polypharmacy across multiple diseases. In each instance, the performance comparison is visualized using box plot. Each box plot shows the distribution of the model’s performance on the 10-fold data associated with the respective cohort. The average RMSE score is denoted by the red triangle and is also annotated as μ. The median score is indicated by the green horizontal line and the standard deviation of the 10-fold scores is denoted by s. We also perform a Student’s t-test to highlight the difference in performance through the computation of p-value (ρ).

First, as depicted in Figure 6, among the five HF drug categories, the model’s ability to predict the physiological response of the patients taking medication under the Beta Blocker category is relatively better with statistical significance compared to the remaining four categories. The bar chart in the Figure 5 subplot shows the most frequently taken medications under each category in our dataset. We consider only the top medication per category, that is the one with the most # of patients on the y-axis and compare their performances in Figure 7. The top drug from Beta Blocker, namely Metoprolol, performs the best followed by Losartan from class ARB. Then moving to drug combinations, the Figure 5 subplot shows the most frequent drug combinations within each category in our dataset. To evaluate the intra-category performance, we only include the top four drug combinations across all the categories for analysis as the other combinations had fewer patient instances for the performance to be reflective of the whole cohort. As depicted in Figure 8, the ARB drug combination comprising Losartan and Valsartan medications gave the best performance with around 42% improvement ahead of the second-best combination, Pravastatin and Simvastatin, belonging to the category Statin. Figure 5 subplot shows the inter-category drug combination frequencies for the top ten combinations found in our dataset. On comparing their performances in Figure 9, the combination of the medications Metoprolol and Furosemide from the categories Beta Blocker and Loop diuretic, respectively, performed the best by reducing the RMSE by around 13% compared to the second-best drug combination Metoprolol from Beta Blocker and Atorvastatin from Statin. To demonstrate the case of polypharmacy across multiple diseases, we consider the medications taken for diabetes mellitus along with the heart failure medications. Figure 5 subplot shows the frequency of the top drug combinations for the two diseases found in our dataset. As shown in Figure 10, the HF drug Metoprolol and diabetes mellitus drug Metformin in combination performed the best by approximately 4%.

**Figure 5.**
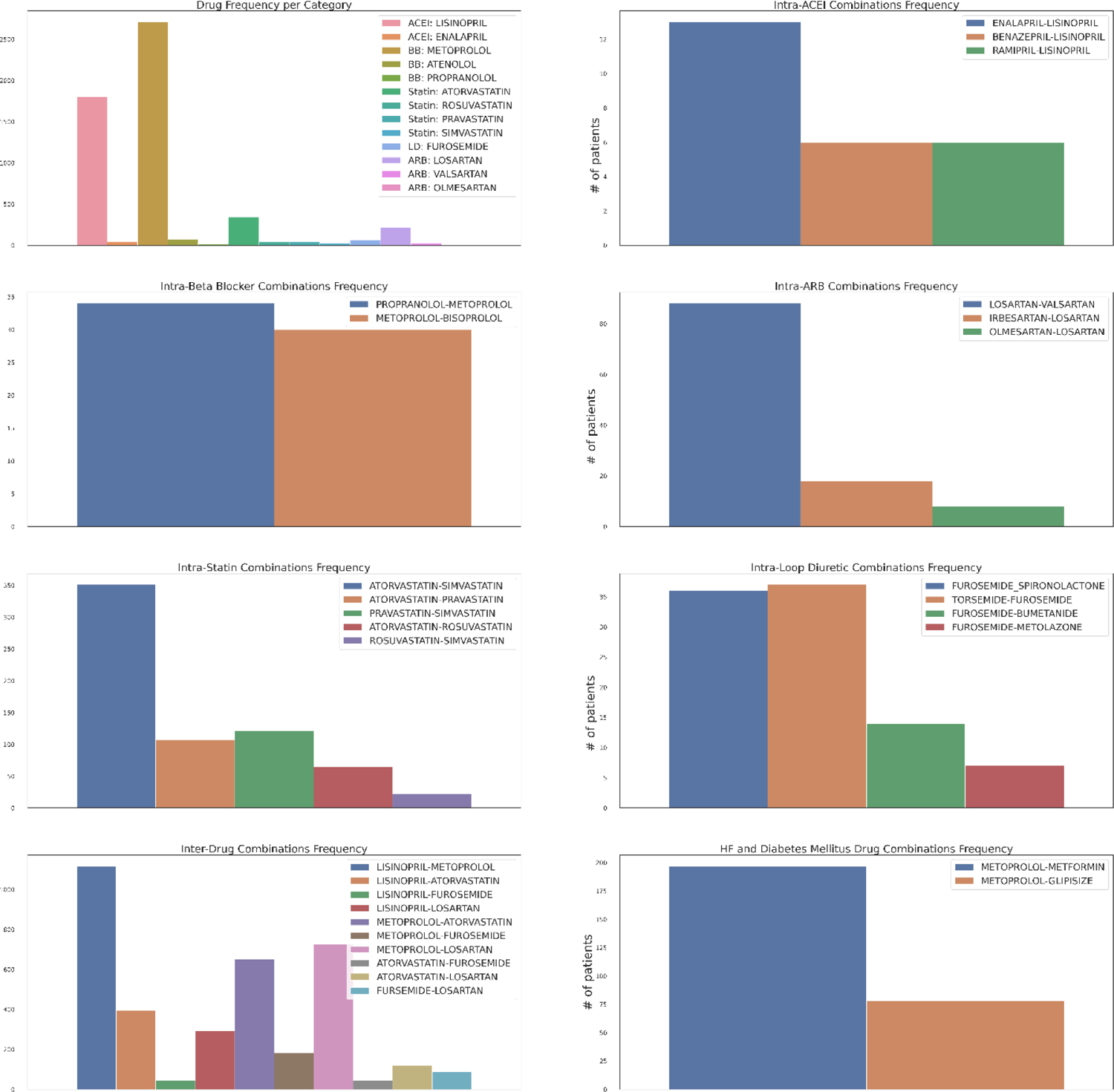
Medication Statistics

**Figure 6.**
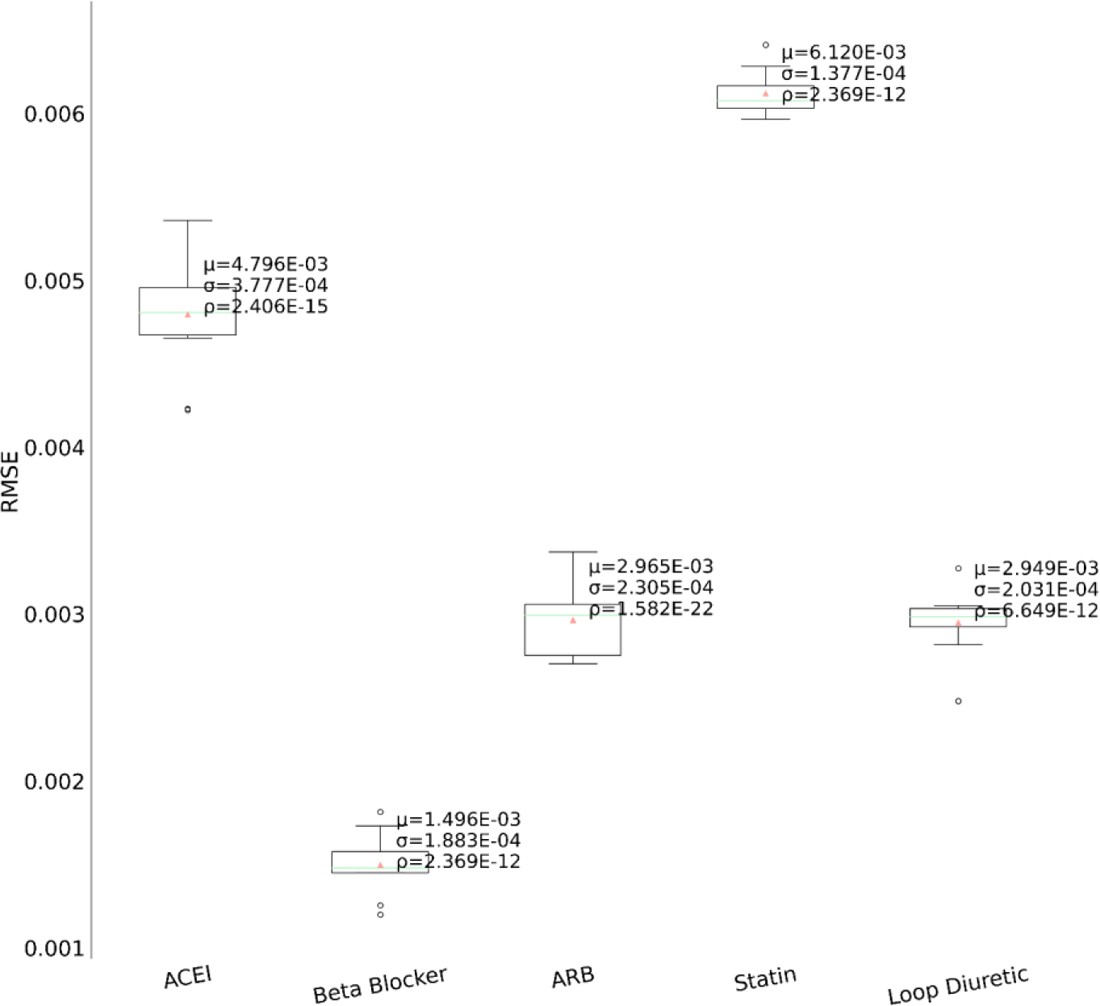
Performance comparison among the medication categories

**Figure 7.**
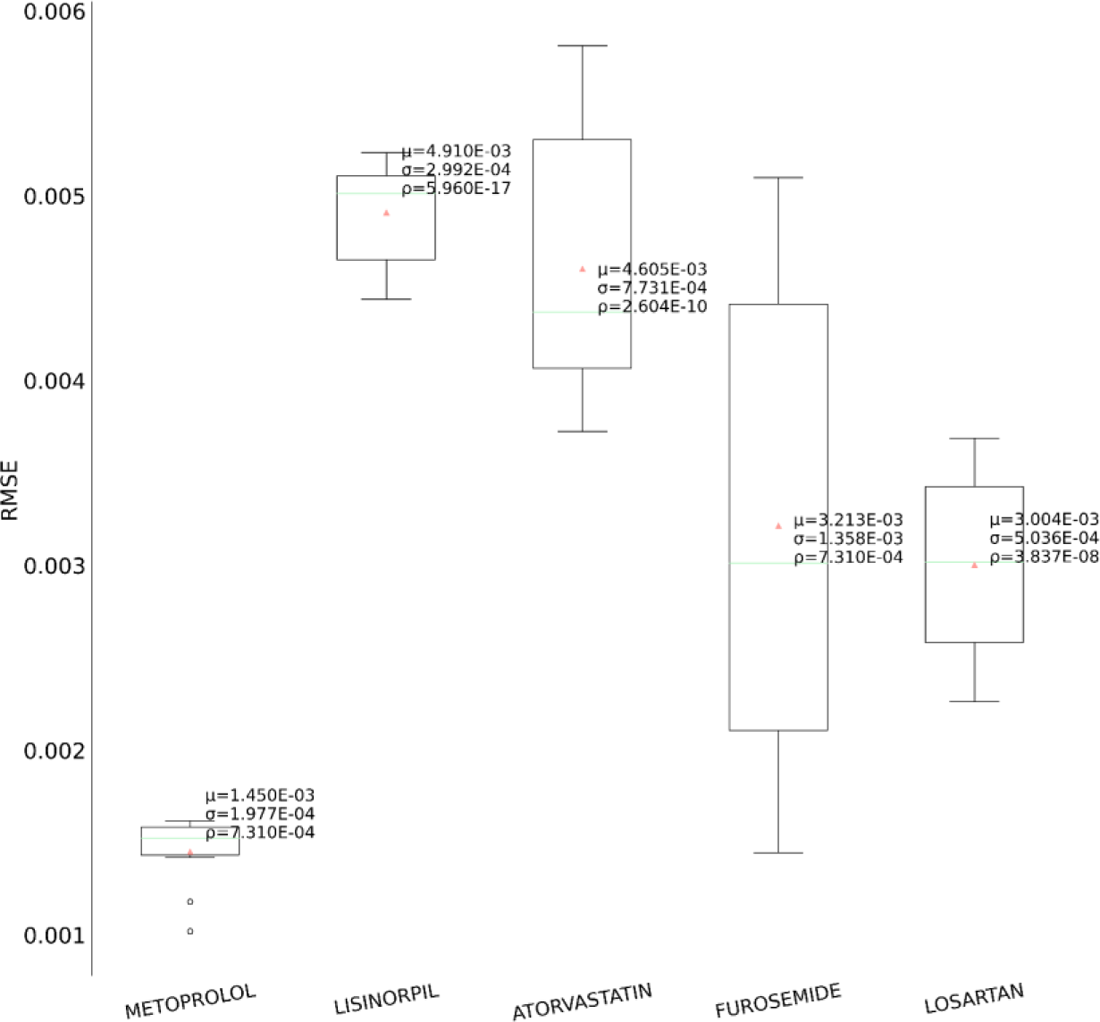
Performance comparison for the top drug within each medication category

**Figure 8.**
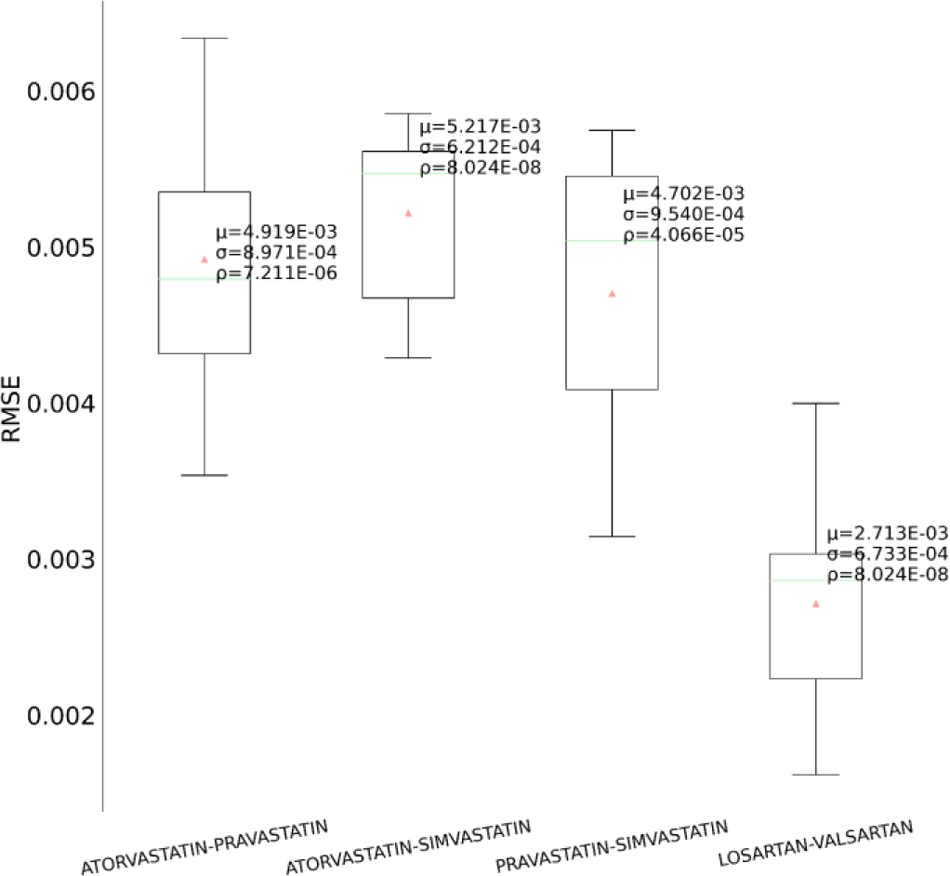
Performance comparison among the intra-category medication combinations

**Figure 9.**
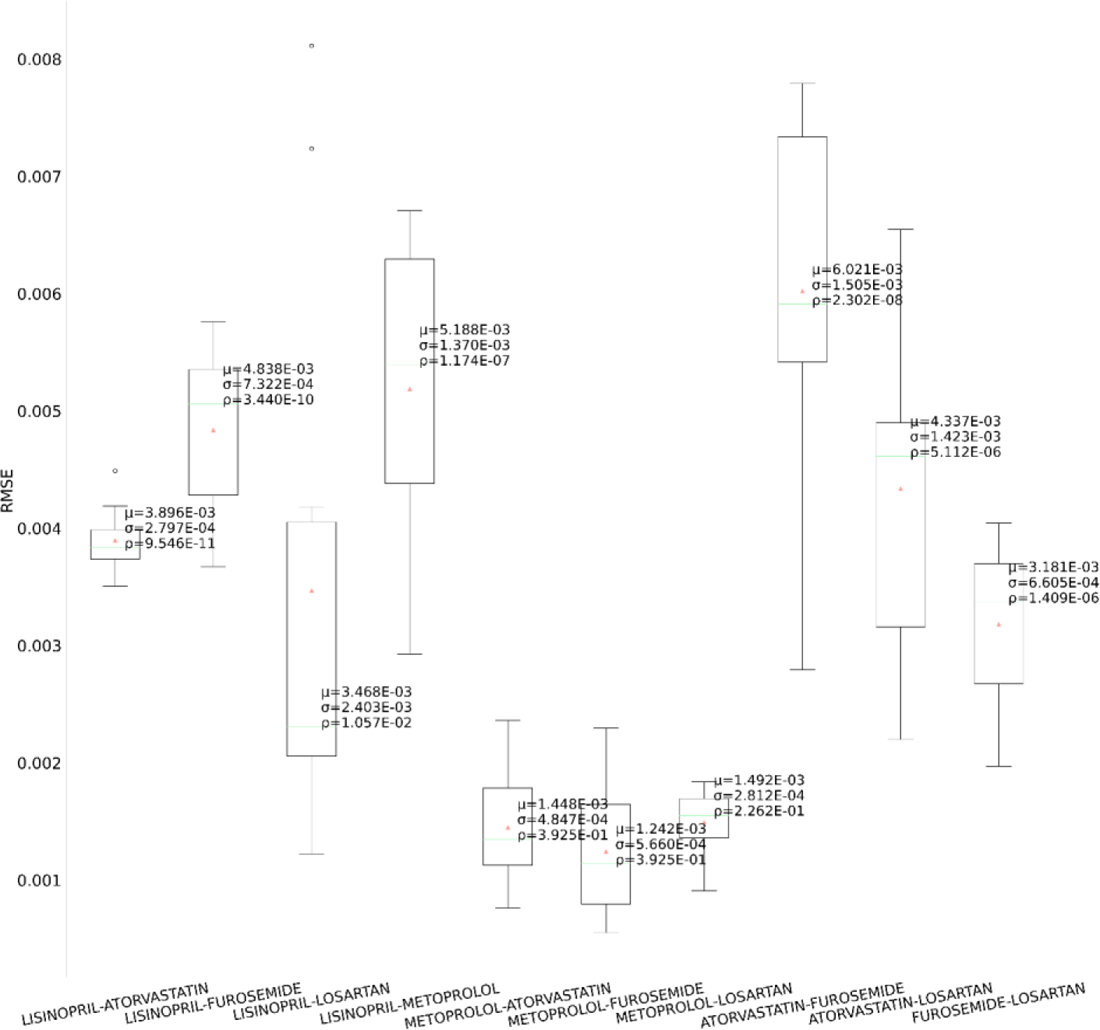
Performance comparison among the inter-category medication combinations

**Figure 10.**
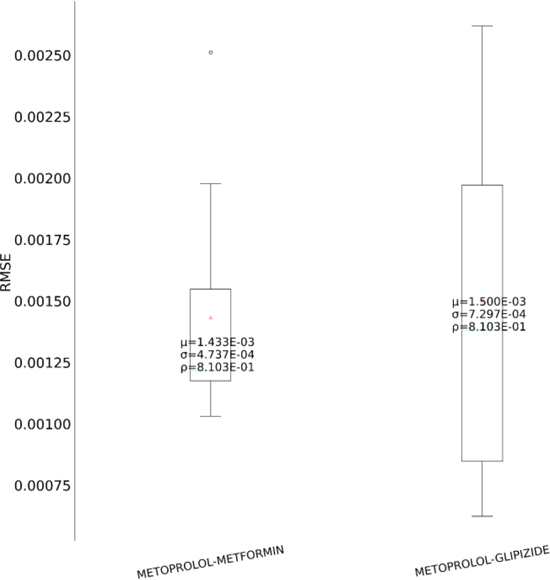
Performance comparison for polypharmacy between HF and diabetes mellitus

### Part 3 Evaluation

Heart failure can be classified according to the left ventricular ejection fraction (LVEF) into two major subtypes - HF with reduced ejection fraction (HFrEF; LVEF ≤ 40%) and HF with preserved ejection fraction (HFpEF; LVEF ≥ 50%)^7^. Figure 11 shows the patient counts in our dataset for the two HF subtypes across the five drug categories. As there exists a dichotomy in the pathophysiology and etiology defining the two subtypes^42^, it would be enlightening to quantify the extent of their treatment response differences to decide effective treatment options. We depict the performance comparison of the model’s generalizability on three cohorts comprised of - HFrEF patients, HFpEF patients and both, as shown in Figure 12. Surprisingly, although HFpEF is considered to be more heterogeneous and resistant to conventional drug therapies^43^, it performs better than HFrEF by ∼31%. This cements the utility of the longitudinal phenotypic features in the EHR as an indispensable resource for heterogeneous treatment analysis.

**Figure 11.**
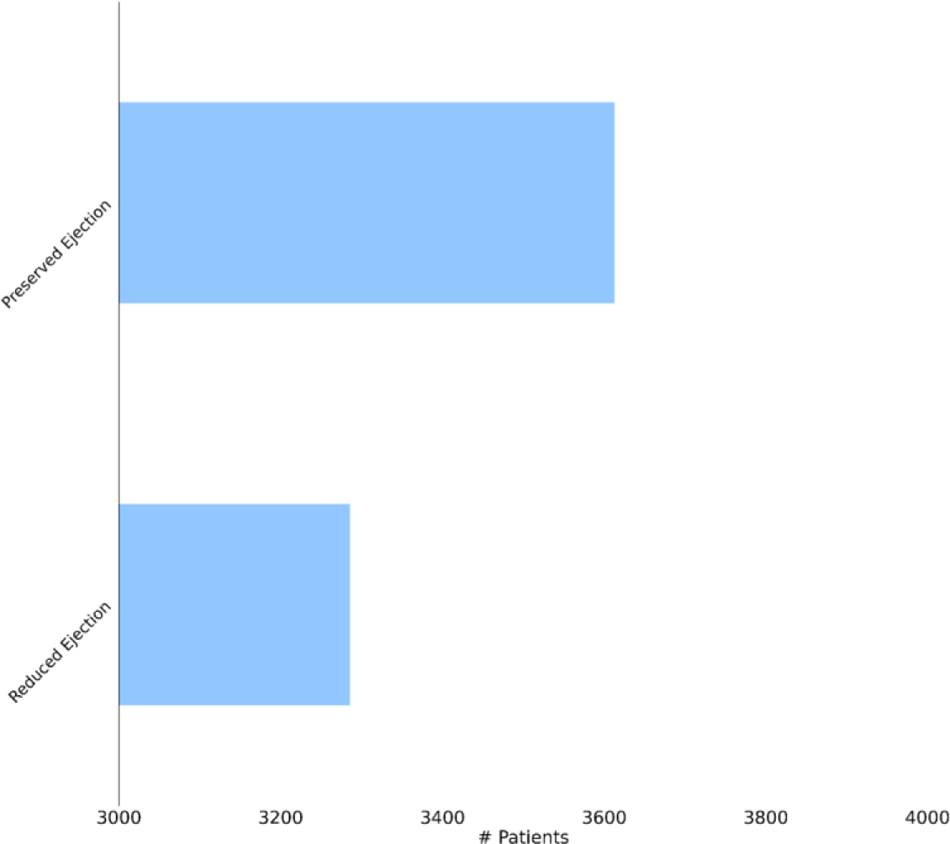
Patient counts from our dataset for the HF subtypes

**Figure 12.**
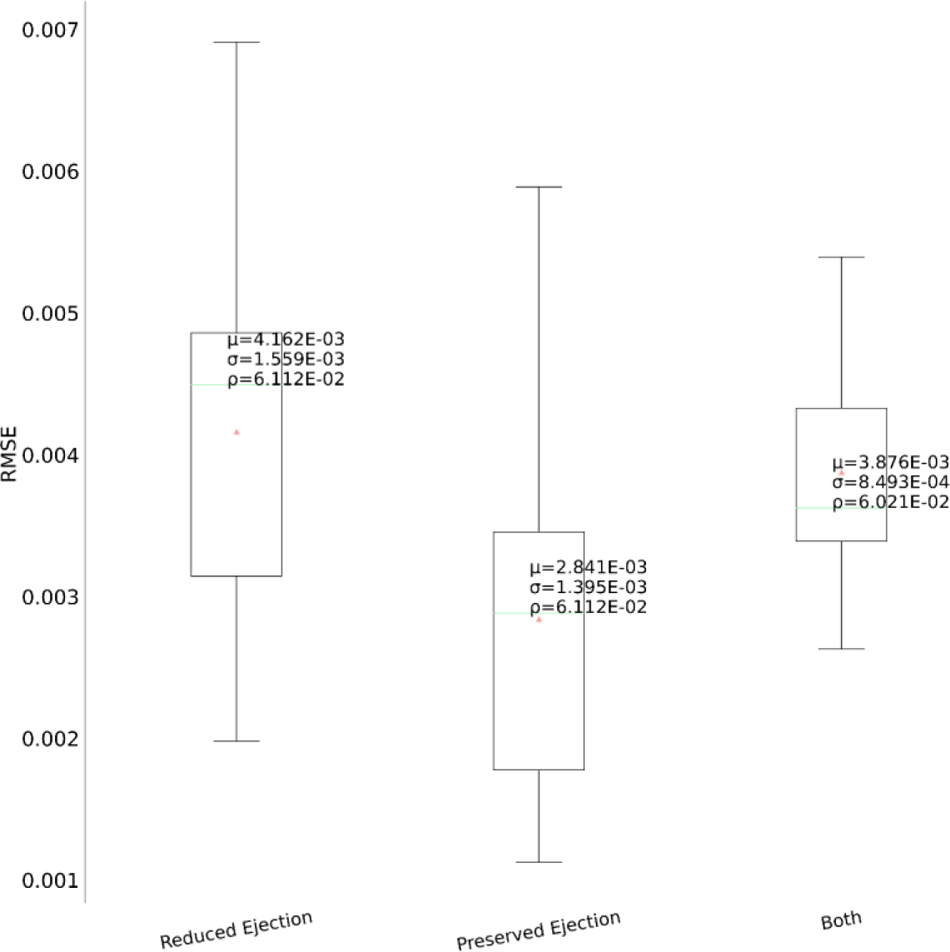
Performance comparison between the HF subtypes

We also provide an intrinsic evaluation by projecting the learned patient representations in a low dimensional space using t-SNE^44^, depicted in Figure 13. The idea is that patients belonging to the same subtype would have similar representations, so would be grouped together. The representation strength of our graph-based framework substantiates this as patients have been separated into two distinct clusters corresponding to the two HF subtypes.

**Figure 13.**
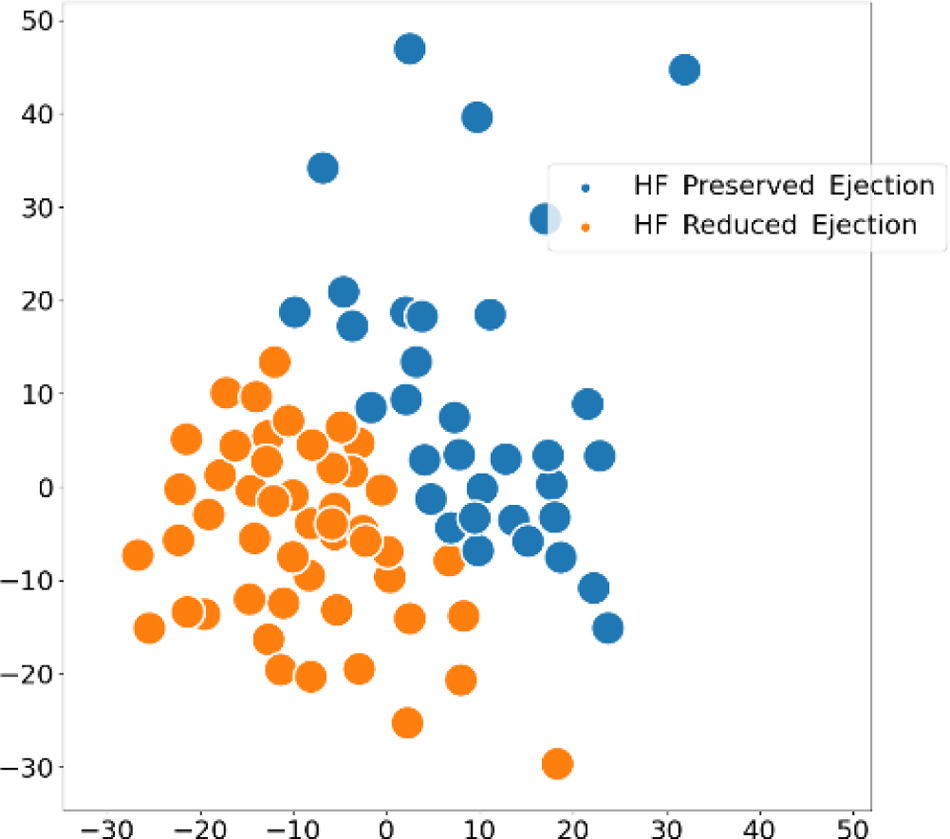
t-SNE visualization of the learned embedding space

## Conclusion

In this work, we introduce a novel graph-based framework for HF treatment outcome prediction. Our study demonstrates that it is possible to effectively forecast the patient’s physiological response in the future visit by modeling the spatial-temporal correlations in the heterogeneous EHR observations as graph-structured data. We validate the superiority of our framework rigorously through a series of experiments on a real-world clinical data and evaluate using three error metrics.

## Data Availability

All data used in the present study are PHI so cannot be made publicly available.

## Acknowledgements

This study is supported by the National Institute of Health (NIH) NIGMS (R00GM135488).

